# Patient Insights into the Diagnosis of Smell and Taste Disorders in the United States

**DOI:** 10.1101/2023.09.20.23295861

**Authors:** Bita R. Naimi, Stephanie R. Hunter, Katie Boateng, Nancy E. Rawson, Emily Garvey, Pamela H. Dalton, Jenifer Trachtman, Claire Murphy, Paule V. Joseph, Suz Schrandt, Pamela Silberman, Alexander Duffy, Gurston G. Nyquist

**Affiliations:** Department of Otolaryngology, Thomas Jefferson University Hospital, Philadelphia PA USA; Monell Chemical Senses Center, Philadelphia PA USA; Smell and Taste Association of North America, Philadelphia PA USA; Department of Psychology, San Diego State University, San Diego CA USA; National Institute of Alcohol Abuse and Alcoholism and National Institute of Nursing Research, Section of Sensory Science and Metabolism, Bethesda MD USA; ExPPect, LLC, Arlington VA USA

**Keywords:** olfaction, olfactory disorders, olfactory test, quality of life

## Abstract

**OBJECTIVES:** Diagnosis of smell/taste dysfunction is necessary for appropriate medical care. This study examines factors affecting testing and diagnosis of smell/taste disorders.

**METHODS:** The online *USA Smell and Taste Patient Survey* was made available to US patients with smell/taste disorders between April 6-20, 2022. 4,728 respondents were included.

**RESULTS:** 1,791 (38%) patients reported a documented diagnosis. Patients most often saw family practitioners (34%), otolaryngologists (20%), and Taste/Smell clinics (6%) for smell/taste dysfunction. 64% of patients who went to Taste/Smell clinics received smell testing, followed by 39% of patients who saw otolaryngologists, and 31% of patients who saw family practitioners. Factors associated with increased odds of diagnosis included age (25-39 years (OR 2.97, 95% CI [2.25, 3.95]), 40-60 (OR 3.3, 95% CI [2.56, 4.52]), and >60 (OR 4.25, 95% CI [3.21, 5.67]) vs. 18-24 years), male gender (OR 1.26, 95% CI [1.07, 1.48]), insurance status (private (OR 1.61, 95% CI [1.15, 2.30]) or public (OR 2.03, 95% CI [1.42, 2.95]) vs. uninsured), perception of their family practitioner to be knowledgeable (OR 2.12, 95% CI [1.16, 3.90]), otolaryngologic evaluation (OR 6.17, 95% CI [5.16, 7.38]), and psychophysical smell testing (OR 1.77, 95% CI [1.42, 2.22]).

**CONCLUSION:** Psychophysical testing, otolaryngologic evaluation, patient assessment of family practitioner knowledge level, insurance, age, and gender are significant factors in obtaining smell/taste dysfunction diagnosis. This study identifies barriers to diagnosis including lack of insurance or access to specialist evaluation and highlights the importance of educating family practitioners in diagnosis and management of patients with smell/taste disorders.

## INTRODUCTION

Prior to the COVID-19 pandemic, an estimated 27 million United States adults experienced olfactory and gustatory disturbances.^1^ The COVID-19 pandemic has resulted in a higher incidence and increased awareness of smell or taste disorders with an estimated 60% of the adult population experiencing smell and taste-related symptoms accompanying COVID-19 infection.^2–6^ Smell and taste disorders encompass many etiologies, including infectious (COVID-19, viral upper respiratory infection), inflammatory (nasal and paranasal sinus disease), trauma, toxins, malignancy, congenital, pregnancy-related changes, neurodegenerative disorders, and idiopathic etiologies.^7–9^ Given the wide range of causative etiologies, diagnosis and management of these disorders may prove challenging. Psychophysical testing for smell or taste dysfunction is crucial to ensure accurate diagnosis and provide adequate care.^10^ However, there have been many anecdotal reports of medical providers failing to hear their patients’ concerns and to provide a clear diagnosis.^11,12^ Patients are often uncertain of their prognosis or next steps in their medical care.

Smell and taste dysfunction has been shown to be detrimental to quality of life.^13–15^ Patients with undiagnosed or untreated disease are especially at risk of significant psychological and physical consequences, including anxiety, depression, malnutrition, and safety concerns.^16,17^ Establishing a diagnosis of smell or taste dysfunction is important to ensure that patients receive appropriate treatment, follow up, and necessary medical and social supports. This study utilized a nationally distributed survey to understand patient- and provider-related factors associated with testing and establishing a diagnosis of smell or taste dysfunction for patients in the United States.

## MATERIALS AND METHODS

This survey study of patients with smell and taste disorders in the United States was granted exemption by the University of Pennsylvania Institutional Review Board under protocol number 851039, through which Monell Chemical Senses Center is affiliated.

### USA Smell and Taste Patient Survey

The USA Smell and Taste Patient Survey **(Supplemental File)** was developed through collaboration between the Department of Otolaryngology at Thomas Jefferson University Hospital, the Smell and Taste Association of North America (STANA), a chemosensory consultant, and the Monell Chemical Senses Center (Philadelphia, PA, USA) to understand the impact of smell or taste dysfunction from COVID-19 and other causes on everyday experiences, and to gain a better understanding of barriers to diagnosis or treatment for smell and taste loss. This 60-question non-incentivized survey was made available online to patients with smell and taste dysfunction, from April 6-20, 2022, distributed through STANA, Monell, and Jefferson email newsletters, and STANA social media pages including Twitter, LinkedIn, Instagram, and Facebook. Survey questions included patient demographics, insurance status, self-reported diagnosis status, onset and etiology of smell/taste dysfunction, diagnostic testing administered, reasons for difficulties in obtaining a diagnosis, and type of provider seen. Patients were also asked to rate their perception on how knowledgeable their provider was about smell or taste dysfunction, ranging from “not knowledgeable at all,” “slightly knowledgeable,” “moderately knowledgeable,” “very knowledgeable,” to “extremely knowledgeable.” In the analysis, perception of provider knowledge was grouped into a ‘low knowledge’ category, consisting of “not knowledgeable at all” and “slightly knowledgeable” responses; and a ‘high knowledge’ category, consisting of “moderately knowledgeable,” “very knowledgeable,” and “extremely knowledgeable” responses. Survey

### Statistical Analysis

Multivariate logistic regression was performed in R (Version 2022.12.0+353) to assess the effect of age, gender, smell testing, symptom onset, insurance status, and visit with an otolaryngologist on having a documented diagnosis of smell and taste dysfunction. Separate multivariate logistic regression models were conducted to assess the effects of each type of testing performed on obtaining a diagnosis, the effects of perceived provider knowledge on obtaining a diagnosis, and the effects of different providers seen on performing smell testing. Age and gender were included in each model. Statistical significance was set at p<0.05. Odds ratios with 95% confidence intervals were calculated for statistically significant factors. Variance inflation factors were used to assess multicollinearity.

## RESULTS

A total of 6,100 participants provided electronic informed consent and completed this survey, of which 5,815 indicated they resided in the United States. 5,528 participants indicated that they have smell and/or taste loss, referred to here as patients. Participants less than 18 years old (n=176), those that did not answer insurance status (n=167), and who did not know if their smell or taste dysfunction was due to COVID-19 (n=604) were excluded. Regarding gender, participants who answered non-binary, prefer not to say, or other (n=34) were excluded due to the small sample size that could not be statistically analyzed. Thus, 4,728 patients with smell or taste dysfunction were included in the analyses. Patient demographics are summarized in **Table 1**; the cohort consisted of 73% females and 27% males, and was predominantly white (88%), insured by private insurance (66%), and aged 40-60 years old (39%).

**Table 1.**
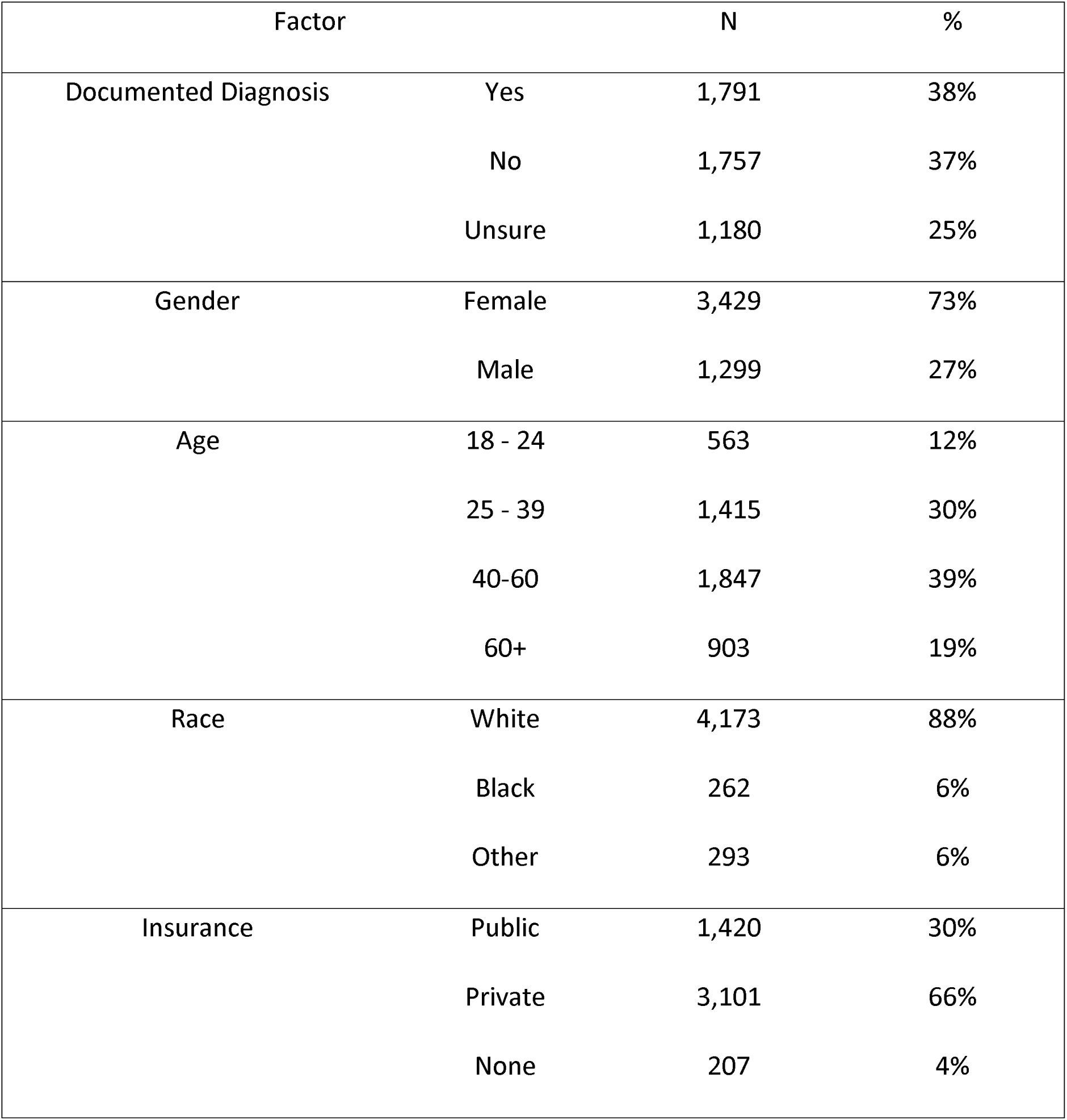
Patient Summary.

| Factor |  | N | % |
| --- | --- | --- | --- |
| Documented Diagnosis | Yes | 1,791 | 38% |
|  | No | 1,757 | 37% |
|  | Unsure | 1,180 | 25% |
| Gender | Female | 3,429 | 73% |
|  | Male | 1,299 | 27% |
| Age | 18 - 24 | 563 | 12% |
|  | 25 - 39 | 1,415 | 30% |
|  | 40-60 | 1,847 | 39% |
|  | 60+ | 903 | 19% |
| Race | White | 4,173 | 88% |
|  | Black | 262 | 6% |
|  | Other | 293 | 6% |
| Insurance | Public | 1,420 | 30% |
|  | Private | 3,101 | 66% |
|  | None | 207 | 4% |

53% of patients in this study sought care from a healthcare provider for their smell or taste dysfunction. The providers seen most often for smell or taste dysfunction were family practitioners (n=1,612 (34%)), followed by otolaryngologists (n=929 (20%)), Taste and Smell clinics (n=262 (6%)), neurologists (n=203 (4%)), and nurse practitioners (n=181 (4%)) (**Table 2**). 1,791 (38%) patients had a diagnosis documented in their medical record, 1,757 (37%) did not have a diagnosis, and 1,180 (25%) were unsure **(Table 1)**. Factors associated with obtaining a diagnosis are summarized in **Table 3**. Patients aged 25-39 (OR 2.97, 95% CI [2.25, 3.95]), 40-60 (OR 3.39, 95% CI [2.56, 4.52]), and >60 (OR 4.25, 95% CI [3.21, 5.67]) were more likely to have a diagnosis than those aged 18-24. Males (OR 1.26, 95% CI [1.07, 1.48]) were more likely to have a diagnosis than females. Patients with private (OR 1.61, 95% CI [1.15, 2.30]) and public (OR 2.03, 95% CI [1.42, 2.95]) health insurance were more likely to have a diagnosis than uninsured patients. Patients who saw an otolaryngologist (OR 6.17, 95% CI [5.16, 7.38]) and who received a smell test (OR 1.77, 95% CI [1.42, 2.22]) were more likely to have a diagnosis than those who did not. Those who had smell or taste dysfunction as a sequela of COVID-19 infection were more likely to have a documented diagnosis than those with smell or taste dysfunction from another etiology (OR 1.13, 95% CI [1.10, 1.58]), and those whose symptoms started after the pandemic were more likely to have a diagnosis than those with pre-pandemic dysfunction (OR 1.23, 95% CI [1.07, 1.40]).

**Table 2.**
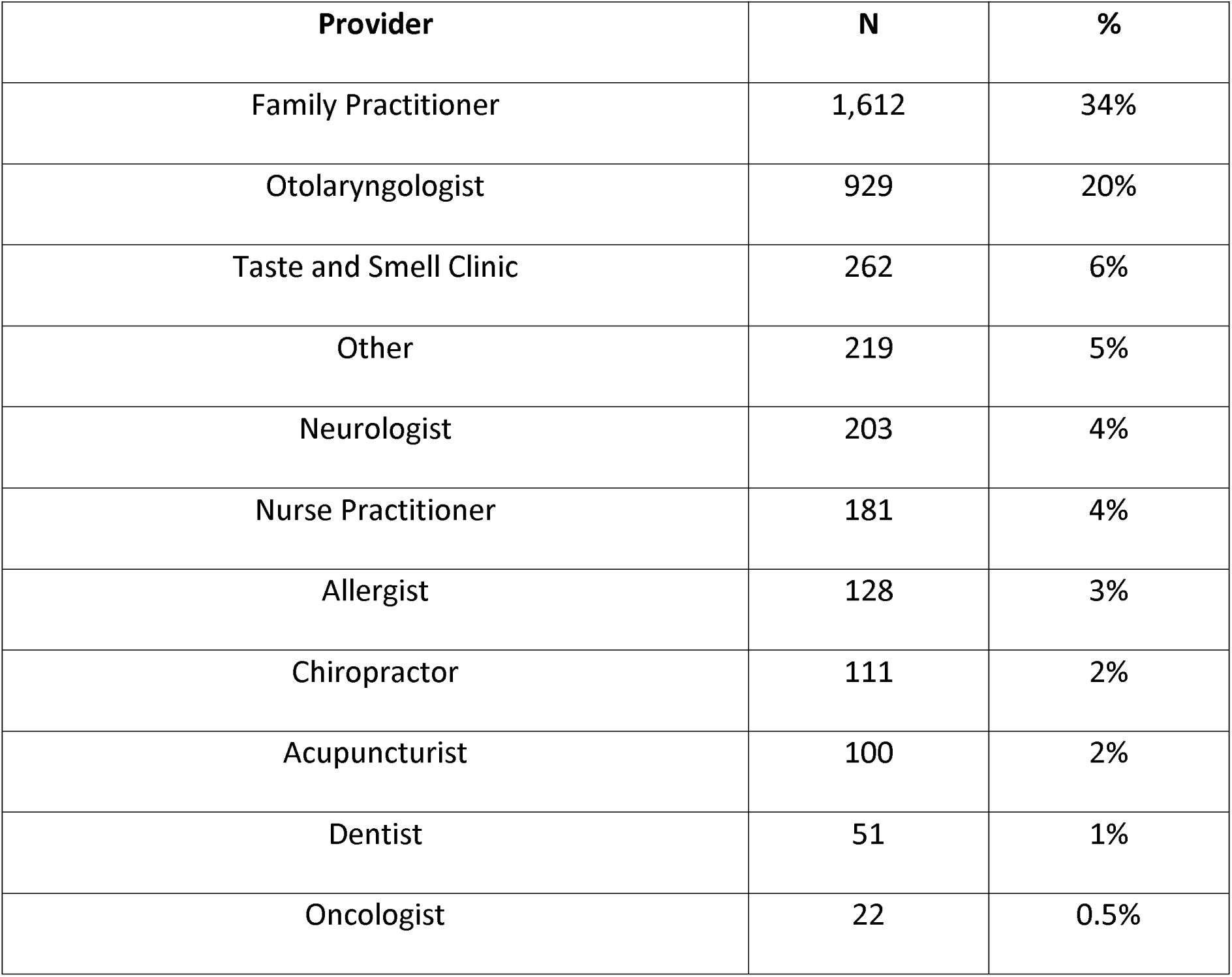
Providers Seen for Smell or Taste Dysfunction.

| **Provider** | **N** | **%** |
| --- | --- | --- |
| Family Practitioner | 1,612 | 34% |
| Otolaryngologist | 929 | 20% |
| Taste and Smell Clinic | 262 | 6% |
| Other | 219 | 5% |
| Neurologist | 203 | 4% |
| Nurse Practitioner | 181 | 4% |
| Allergist | 128 | 3% |
| Chiropractor | 111 | 2% |
| Acupuncturist | 100 | 2% |
| Dentist | 51 | 1% |
| Oncologist | 22 | 0.5% |

**Table 3.**
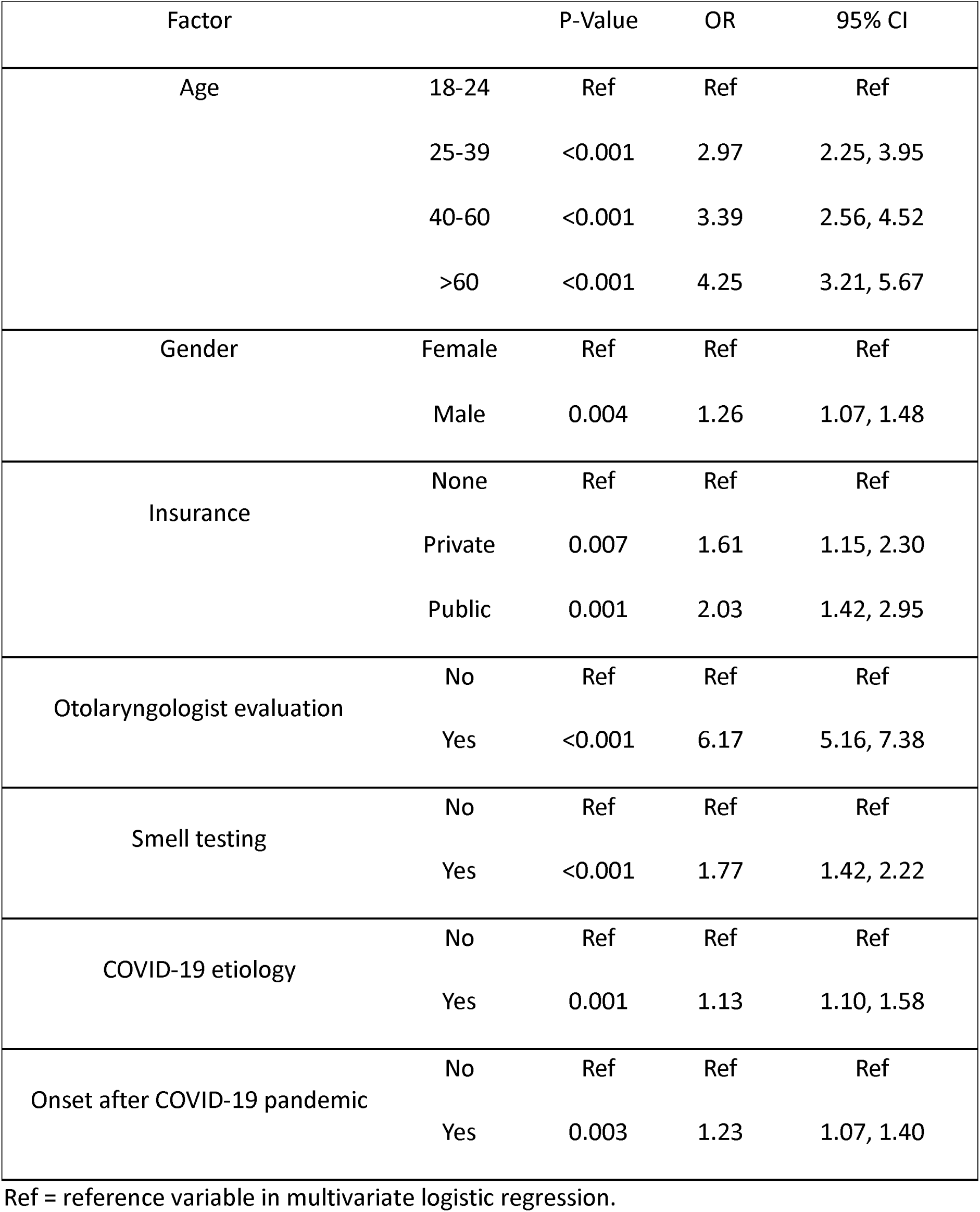
Factors Associated with Smell or Taste Dysfunction Diagnosis.

| Factor |  | P-Value | OR | 95% CI |
| --- | --- | --- | --- | --- |
| Age | 18-24 | Ref | Ref | Ref |
|  | 25-39 | <0.001 | 2.97 | 2.25, 3.95 |
|  | 40-60 | <0.001 | 3.39 | 2.56, 4.52 |
|  | >60 | <0.001 | 4.25 | 3.21, 5.67 |
| Gender | Female | Ref | Ref | Ref |
|  | Male | 0.004 | 1.26 | 1.07, 1.48 |
| Insurance | None | Ref | Ref | Ref |
|  | Private | 0.007 | 1.61 | 1.15, 2.30 |
|  | Public | 0.001 | 2.03 | 1.42, 2.95 |
| Otolaryngologist evaluation | No | Ref | Ref | Ref |
|  | Yes | <0.001 | 6.17 | 5.16, 7.38 |
| Smell testing | No | Ref | Ref | Ref |
|  | Yes | <0.001 | 1.77 | 1.42, 2.22 |
| COVID-19 etiology | No | Ref | Ref | Ref |
|  | Yes | 0.001 | 1.13 | 1.10, 1.58 |
| Onset after COVID-19 pandemic | No | Ref | Ref | Ref |
|  | Yes | 0.003 | 1.23 | 1.07, 1.40 |

Ref = reference variable in multivariate logistic regression.

Patient-reported reasons for difficulties in obtaining a diagnosis are demonstrated in **Figure 1**. This survey question was a “select all that apply” style question, with 2,458 (52%) patients selecting at least one difficulty. The most selected reason was lack of provider knowledge (16%), followed by not being listened to (14%), lack of access to a specialist (5%), symptoms being attributed to another disease (4%), lack of insurance (3%), and cost-prohibitive nature of testing (2%).

**Figure 1.**
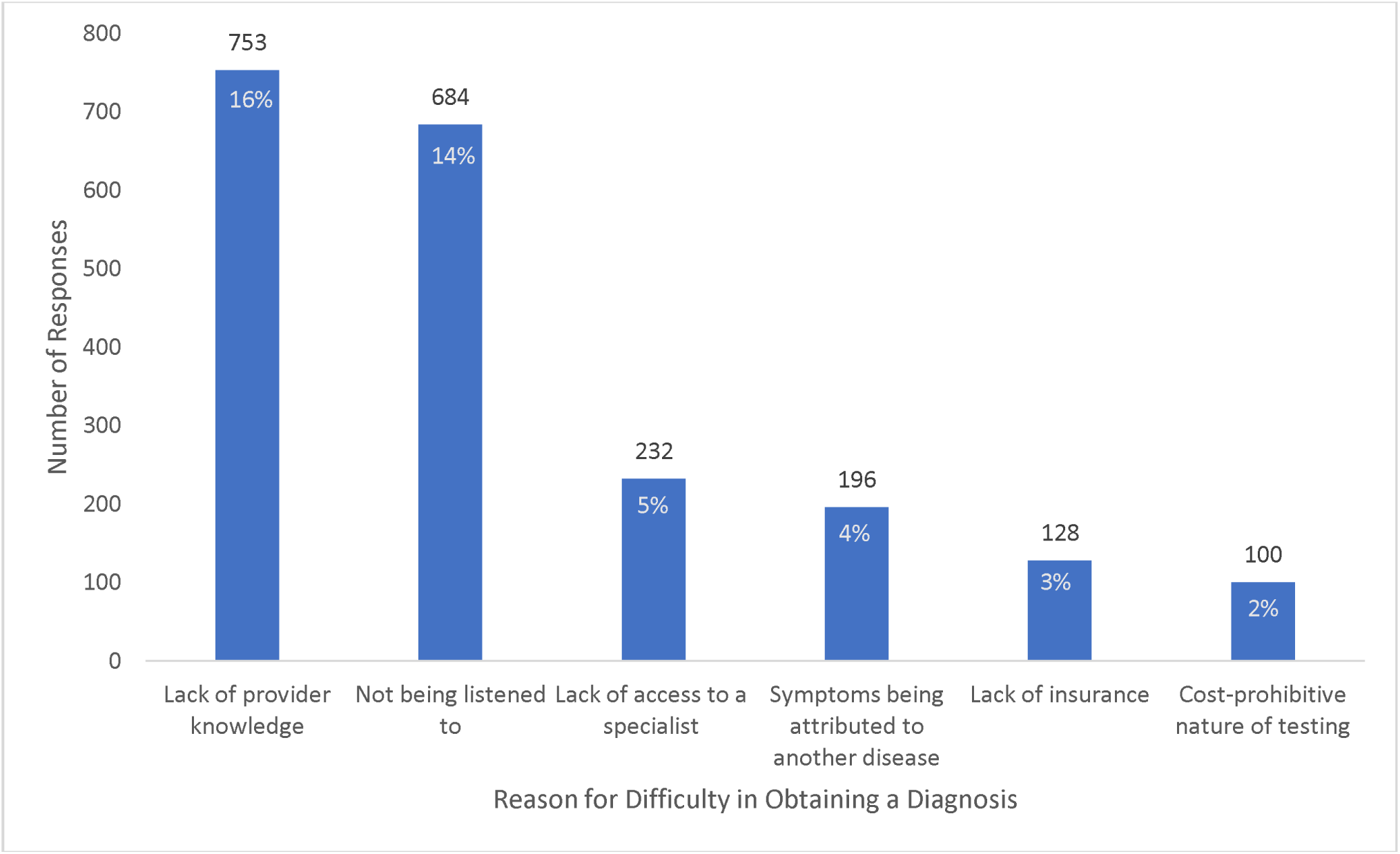
“Select all that apply” responses to why patients had difficulty obtaining a diagnosis for their smell or taste dysfunction. 2,458 (52%) of the patients selected at least one answer.

The impact of perceived provider knowledge on obtaining a diagnosis was assessed for each type of provider seen. Family practitioner knowledge had a significant impact on patient diagnosis (p=0.02) **(Table 4).** Patients who perceived their family practitioner to be knowledgeable about smell or taste dysfunction had significantly greater odds of obtaining a diagnosis (2.12 [1.16, 3.90]), compared to patients who perceived their family practitioner to lack appropriate knowledge. Provider knowledge among otolaryngologists, nurse practitioners, and smell and taste clinics did not have a significant impact on patient diagnosis **(Table 4).**

**Table 4.** Impact of Perceived Provider Knowledge on Smell or Taste Dysfunction Diagnosis.

| Provider Type | P-value | OR | 95% CI |
| --- | --- | --- | --- |
| Otolaryngologist | 0.08 | -- | -- |
| Family Practitioner | 0.02 | 2.12 | 1.16, 3.90 |
| Nurse Practitioner | 0.11 | -- | -- |
| Taste and Smell Clinic | 0.31 | -- | -- |

Patients were asked which diagnostic testing, if any, they underwent; results are summarized in **Figure 2**. This question was a “select all that apply” style question. 18% of patients reported that they did not undergo any testing. 17% underwent psychophysical smell testing, 11% underwent nasal endoscopy, 9% underwent a medical history, 7.6% underwent magnetic resonance imaging (MRI), 6.9% underwent computed tomography (CT) scan, 6.7% underwent taste testing, and 3.6% filled out a questionnaire. Patients had different odds of diagnosis depending on testing, with results summarized in **Table 5**. Patients who underwent taste testing had the highest odds of diagnosis (OR 9.77, 95% CI [7.24, 13.40]), followed by MRI (OR 6.98, 95% CI [5.34, 9.24]), CT (OR 6.73, 95% CI [5.10, 9.01]), and nasal endoscopy (OR 6.69, 95% CI [5.36, 8.42]).

**Figure 2.**
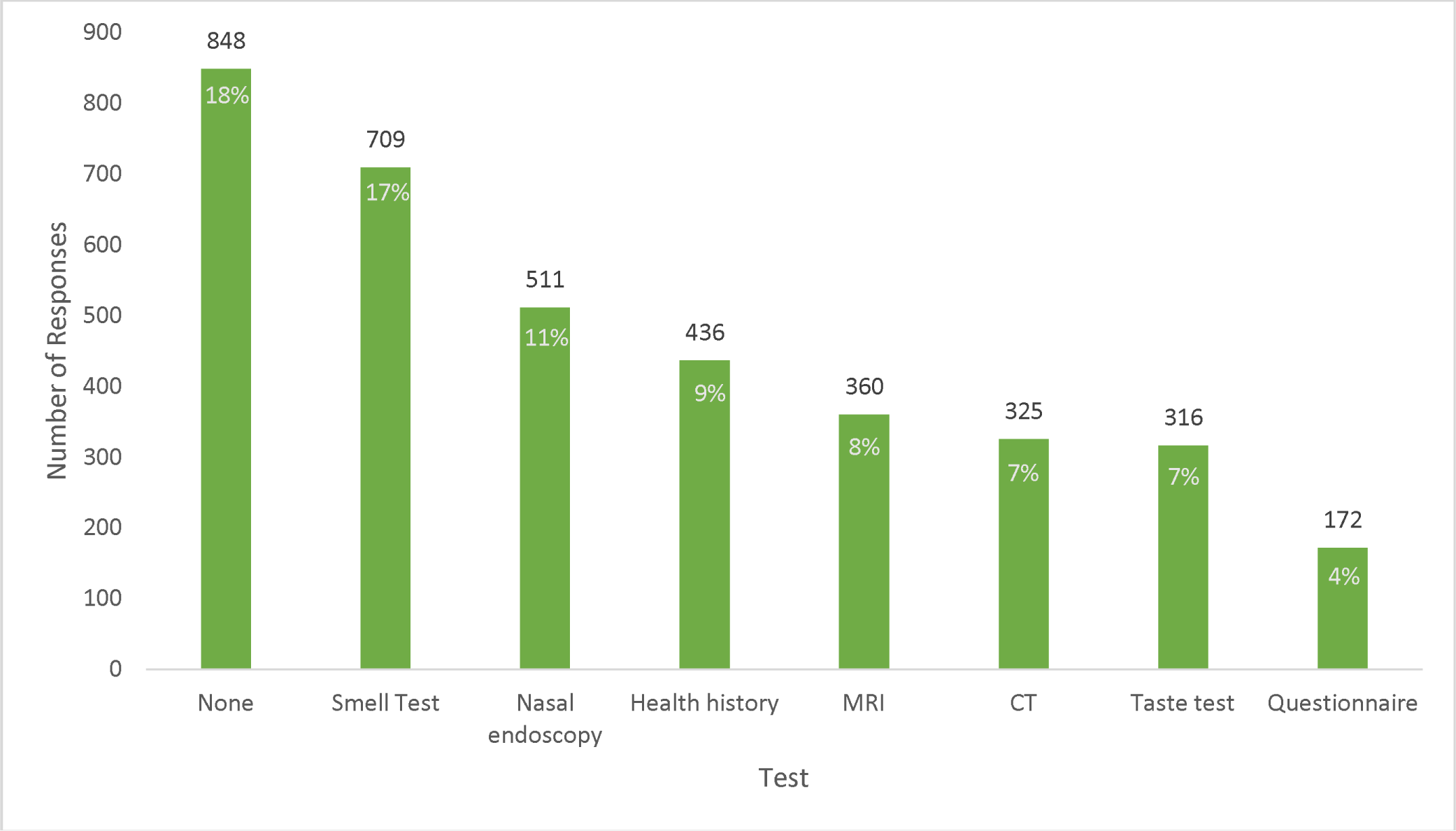
“Select all that apply” responses for diagnostic testing that patients underwent.

**Table 5.** Diagnostic Testing Impact on Smell or taste Dysfunction Diagnosis.

| Test | P-value | OR | 95% CI |
| --- | --- | --- | --- |
| Taste Test | <0.001 | 9.77 | 7.24, 13.40 |
| MRI | <0.001 | 6.98 | 5.34, 9.24 |
| CT | <0.001 | 6.73 | 5.10, 9.01 |
| Endoscopy | <0.001 | 6.69 | 5.36, 8.42 |
| Questionnaire | <0.001 | 6.43 | 4.45, 9.54 |
| History | <0.001 | 5.67 | 4.50, 7.21 |
| Smell Test | <0.001 | 3.28 | 2.71, 3.98 |
| Multiple Tests | <0.001 | 2.69 | 2.49, 2.92 |

Among the different types of diagnostic testing available, the frequency of smell testing was analyzed. **Table 6** summarizes the proportion of patients who obtained smell testing based on the type of provider seen for their smell or taste dysfunction. 64% of patients who went to a Taste and Smell clinic obtained smell testing (OR 10.60 [95% CI 7.87, 14.35]), followed by 39% of patients who saw otolaryngologists (OR 10.24 [CI 8.3, 12.70]), 34% of patients who saw neurologists (OR 4.25 [CI 3.05, 5.89]), and 31% of patients who saw family practitioners (OR 3.42 [CI 2.87, 4.09]).

**Table 6.** Smell Testing Based on Provider Seen for Smell or Taste Dysfunction.

| Provider | Smell Testing Performed |  | OR | 95% CI |
| --- | --- | --- | --- | --- |
|  | Yes | No |  |  |
| Taste and Smell Clinic | 181 (64%) | 103 (36%) | 10.60 | 7.87, 14.35 |
| Otolaryngologist | 366 (39%) | 563 (61%) | 10.24 | 8.28, 12.70 |
| Allergist | 49 (38%) | 79 (62%) | 4.70 | 3.14, 6.97 |
| Neurologist | 70 (34%) | 133 (66%) | 4.25 | 3.05, 5.89 |
| Acupuncture | 33 (33%) | 67 (67%) | 4.16 | 2.60, 6.56 |
| Family Practitioner | 493 (31%) | 1,119 (70%) | 3.42 | 2.87, 4.09 |
| Chiropractor | 23 (21%) | 88 (79%) | 1.96 | 1.15, 3.22 |
| Other | 25 (11%) | 194 (89%) | 0.99 | 0.62, 1.51 |
| Dentist | 10 (20%) | 41 (80%) | 1.31 | 0.59, 2.67 |
| Nurse Practitioner | 19 (10%) | 162 (90%) | 0.64 | 0.37, 1.06 |
| Oncologist | 1 (5%) | 21 (95%) | 0.17 | 0.01, 0.88 |

## DISCUSSION

Accurate documentation and diagnostic coding is a critical aspect of patient care,^18–21^ as diagnostic coding captures the patient’s state of disease and can be used as a marker over time to track disease progress and treatment efficacy.^21^ Complete documentation is important for ensuring seamless communication about patients’ medical and social needs from provider to provider.^20,21^ With a multitude of etiologies resulting in smell and taste dysfunction, identifying and coding for the appropriate cause is paramount to counseling and subsequent treatment.

In this study, of the 4,728 included survey participants with self-reported smell or taste dysfunction, less than half (38%) reported a documented diagnosis in their medical record. Objective smell testing, diagnostic imaging, evaluation by otolaryngology, and patient perceptions of family practitioner knowledge were statistically significant factors in obtaining smell or taste dysfunction diagnosis.

Proper documentation of smell or taste dysfunction is necessary to provide patients the appropriate medical and social supports. Studies have shown that patients with olfactory dysfunction experience increased risk of morbidity and mortality compared to their normosmic counterparts, independent of confounding factors such as nutritional status, cognitive function, mental health, smoking and alcohol abuse, and frailty.^22^ Smell and taste disturbances have also been shown to have significant psychosocial implications on patients’ quality of life and well-being, putting patients at higher risk of anxiety and depression.^13,16,17^ Previous literature has demonstrated that worse olfactory function correlates with worse olfactory-specific quality of life.^15^ Patients report the inability to enjoy food, maintain their appetite and nutritional status, participate in social gatherings, and maintain personal hygiene.^13^ Additionally, patients with diminished senses face safety concerns such as the inability to detect smoke, fires, or gas leaks, and can also impact professional careers for chefs, firefighters, and perfumers for example.^13,23,24^ Without documentation and record-keeping of smell or taste dysfunction, new providers may not be aware of the impact of untreated disease on quality of life, nutrition, and physical and mental health.^13^

Olfactory dysfunction can also be a clinical indicator for other disease processes such as neurodegenerative disease or tumors, in which a timely diagnosis is essential.^25^ For example, olfactory dysfunction may be a heralding sign of Parkinson’s disease and can serve as a specific diagnostic marker in the prodromal period.^26^ If olfactory dysfunction is not documented in a patient’s medical record, the provider may not be as diligent in monitoring the patient for early signs of cognitive decline at future visits, and this important clinical information would not be communicated to other treating providers.

Objective smell testing is clinically proven and medically necessary to establish a diagnosis of olfactory dysfunction, and can monitor response to treatments throughout follow up.^24,27,28^ In this study, patients that underwent smell testing were significantly more likely to obtain a diagnosis than those who did not. In this analysis, patients reported undergoing smell testing most often at specialty Taste and Smell clinics, which typically do not accept insurance and cost upwards of $500 for a consultation.^29^ Access to such clinics is limited, as only 6% of participants received care in these settings. Patients had greater access to family practitioners (34%) and otolaryngologists (20%) for evaluation of smell or taste dysfunction. However, only 39% of patients who saw otolaryngologists underwent smell testing, and even fewer patients who saw family practitioners (31%). It is important to note that although family practitioners were the most visited provider for smell or taste dysfunction, less than a third performed smell testing, therefore representing a large, potential cohort of undiagnosed patients.

Outside of self-pay Taste and Smell clinics, medical providers may be less inclined to perform smell testing on patients with smell or taste dysfunction due to lack of insurance reimbursement for such tests^30^. To date, there is no Current Procedural Terminology (CPT) code specifically for olfactory testing, and providers are left with no option but to use the unlisted code 92700 for “unlisted otorhinolaryngological service or procedure.”^30^ This CPT code is often not reimbursed^30,31^. The findings of this study demonstrate the importance of objective smell testing in establishing a diagnosis of olfactory disorders. In addition, our findings support the need for efforts to create a new CPT code and obtain insurance reimbursement of objective psychophysical testing for olfactory dysfunction and other chemosensory disorders.

There are a variety of validated, objective psychophysical tests available for assessing olfactory dysfunction. These include the 40-item Smell Identification Test (UPSIT), the 12-item Brief Smell Identification Test (BSIT), and the 9-item NIH Toolbox Odor Identification Test, which are multiple choice questions presented as microencapsulated odorant strips in a “scratch and sniff” format.^32–36^ These tests measure odor identification, but not threshold or intensity.^32^ These tests are relatively inexpensive, ranging from $20 to $30 per test for the BSIT and UPSIT, and about $2.50 per test for the NIH Toolbox.^33–36^ In order to test odor threshold and discrimination in addition to identification, the Sniffin’ Sticks test may be used.^37–40^ This test is reusable but is significantly more expensive than the “scratch and sniff” format tests, costing approximately $1,325 per set. Sniffin’ Sticks can be burdensome to providers as they are time-consuming to administer and require training for proper administration. Rapid, inexpensive smell tests that can be used as screening tools are being developed but their implementation in clinical practice is still dependent on validation and reimbursement.^41–43^.

The findings of this study demonstrate the importance of provider knowledge on smell or taste dysfunction in establishing a documented diagnosis. Perceived lack of provider knowledge was the most reported reason for why patients experienced difficulty in obtaining a diagnosis. This was demonstrated to be especially impactful within family practitioners, where patients had twice higher odds of receiving a diagnosis if they were seen by a family practitioner perceived to have higher knowledge. This study also demonstrated the importance of specialist evaluation, as patients who saw an otolaryngologist were six times more likely to have received a diagnosis compared to patients who did not see an otolaryngologist. Importantly, family practitioners are often the patient’s first point of contact when encountering the healthcare system, and their awareness and knowledge of smell or taste dysfunction is critical to ensuring the patient receives adequate workup, treatment, follow up, and referral to specialist care when necessary. Providing continuing education for family practitioners on smell or taste dysfunction is especially important given these findings, as the medical field is rapidly evolving with an increased focus on investigation into new experimental treatments for smell or taste dysfunction since the COVID-19 pandemic.^44–46^

Taste testing was the diagnostic modality with the highest odds of obtaining a diagnosis, which could reflect patients who went to a specialist, as 67% of patients who saw a Taste and Smell clinic received taste testing. Patients likely would not have gone to a Taste and Smell Clinic without having or seeking a diagnosis. Diagnostic imaging is also essential for smell or taste dysfunction diagnosis and workup. MRI and CT scan can assess for head and neck pathology that may be causing olfactory disturbance, such as tumors, paranasal sinus disease, and neurodegenerative disease.^47–49^ In this study, various types of diagnostic imaging including CT, MRI, and nasal endoscopy were associated with higher odds of obtaining a documented diagnosis. The odds ratios for CT, MRI, and nasal endoscopy may be high given that providers are required to associate a diagnostic code to order these tests. While imaging is only indicated in select clinical scenarios, these findings highlight the complementary role of imaging to objective olfactory diagnostic testing in the full medical workup for smell or taste dysfunction and in establishing a documented diagnosis.

This study faces several limitations. Although this effort represents one of the largest surveys conducted in those with smell or taste dysfunction in the United States, respondents were mostly white females, limiting the generalizability of results to other gender identities as well as racial and ethnic groups. Second, all responses were self-reported, which inherently limits the certainty of the results, and introduces recall bias. Third, while we asked separate questions about whether patients had a diagnosis documented in their medical record, type of provider seen for smell or taste dysfunction, and type of testing patients underwent, we did not ask which provider recommended the test, or established the diagnosis. Many patients indicated that they saw multiple providers for their smell or taste dysfunction, any of whom could have provided testing or a diagnosis. Nevertheless, this study provides important insights into diagnosis of smell or taste dysfunction.

## CONCLUSION

Objective smell testing, otolaryngologist evaluation, patient perceptions of family practitioner knowledge, insurance, age, and gender are significant factors in obtaining a diagnosis of smell or taste dysfunction. This study highlights the paucity of formal diagnosis of olfactory dysfunction among respondents, and the importance of smell testing and continuing education resources for family practitioners regarding management of these patients. The establishment of a specific CPT code for smell test reimbursement is needed to increase access and utilization of diagnostic objective psychophysical tests. Referral for otolaryngology subspecialty evaluation may prove helpful in obtaining a formal diagnosis of smell and taste dysfunction, as primary care providers may lack extensive knowledge in appropriate diagnostic procedures. Proper documentation is necessary to provide patients the appropriate medical and social support for their safety and in treating smell or taste dysfunction. Lastly, additional patient-participatory studies are needed in the chemosensory field to further understand the impact these smell and taste disorders have on quality of life.

## Supporting information

PCORI Survey Questions

## Data Availability

All data produced in the present study are available upon reasonable request to the authors

## ACKNOWLEDGEMENTS

We thank all the survey respondents for sharing their perspectives and experiences.

## Notes

Financial Disclosures: This work is supported by PCORI Engagement Contract EASC #00261 to NR and NIH grant #R01AG00408526 from the National Institute on Aging to CM. PVJ is supported by the Division of Intramural Research National Institute on Alcohol Abuse and Alcoholism (Z01AA000135), Institute of Nursing Research and the Office of Workforce Diversity, National Institutes of Health Distinguished Scholar, and the Rockefeller University Heilbrunn Nurse Scholar Award.

### Competing Interest Statement

The authors have declared no competing interest.

### Funding Statement

This work is supported by PCORI Engagement Contract EASC #00261 to
NR and NIH grant #R01AG00408526 from the National Institute on Aging to CM. PVJ is
supported by the Division of Intramural Research National Institute on Alcohol Abuse and
Alcoholism (Z01AA000135), Institute of Nursing Research and the Office of Workforce Diversity,
National Institutes of Health Distinguished Scholar, and the Rockefeller University Heilbrunn
Nurse Scholar Award.

## REFERENCES

1. Liu G, Zong G, Doty RL, Sun Q. Prevalence and risk factors of taste and smell impairment in a nationwide representative sample of the US population: a cross-sectional study. BMJ Open. 2016;6(11):e013246. doi:10.1136/bmjopen-2016-013246

2. Aziz M, Perisetti A, Lee-Smith WM, Gajendran M, Bansal P, Goyal H. Taste Changes (Dysgeusia) in COVID-19: A Systematic Review and Meta-analysis. Gastroenterology. 2020;159(3):1132–1133. doi:10.1053/j.gastro.2020.05.003

3. Aziz M, Goyal H, Haghbin H, Lee-Smith WM, Gajendran M, Perisetti A. The Association of “Loss of Smell” to COVID-19: A Systematic Review and Meta-Analysis. Am J Med Sci. 2021;361(2):216–225. doi:10.1016/j.amjms.2020.09.017

4. Agyeman AA, Chin KL, Landersdorfer CB, Liew D, Ofori-Asenso R. Smell and Taste Dysfunction in Patients With COVID-19: A Systematic Review and Meta-analysis. Mayo Clin Proc. 2020;95(8):1621–1631. doi:10.1016/j.mayocp.2020.05.030

5. Xydakis MS, Dehgani-Mobaraki P, Holbrook EH, et al. Smell and taste dysfunction in patients with COVID-19. Lancet Infect Dis. 2020;20(9):1015–1016. doi:10.1016/S1473-3099(20)30293-0

6. Mitchell MB, Workman AD, Rathi VK, Bhattacharyya N. Smell and Taste Loss Associated with COVID-19 Infection. The Laryngoscope. 2023;133(9):2357–2361. doi:10.1002/lary.30802

7. Spielman AI. Chemosensory function and dysfunction. Crit Rev Oral Biol Med Off Publ Am Assoc Oral Biol. 1998;9(3):267–291. doi:10.1177/10454411980090030201

8. Deems DA, Doty RL, Settle RG, et al. Smell and Taste Disorders, A Study of 750 Patients From the University of Pennsylvania Smell and Taste Center. Arch Otolaryngol Neck Surg. 1991;117(5):519–528. doi:10.1001/archotol.1991.01870170065015

9. Harris R, Davidson TM, Murphy C, Gilbert PE, Chen M. Clinical Evaluation and Symptoms of Chemosensory Impairment: One Thousand Consecutive Cases from the Nasal Dysfunction Clinic in San Diego. Am J Rhinol. 2006;20(1):101–108. doi:10.1177/194589240602000119

10. Hannum ME, Ramirez VA, Lipson SJ, et al. Objective Sensory Testing Methods Reveal a Higher Prevalence of Olfactory Loss in COVID-19-Positive Patients Compared to Subjective Methods: A Systematic Review and Meta-Analysis. Chem Senses. 2020;45(9):865–874. doi:10.1093/chemse/bjaa064

11. Hannah’s Anosmia Story. Fifth Sense. Accessed July 7, 2023. https://www.fifthsense.org.uk/stories/hannahs-story/

12. Stories. Fifth Sense. Accessed July 7, 2023. https://www.fifthsense.org.uk/stories/

13. Javed N, Ijaz Z, Khair AH, et al. COVID-19 loss of taste and smell: potential psychological repercussions. Pan Afr Med J. 2022;43:38. doi:10.11604/pamj.2022.43.38.31329

14. Liu DT, Prem B, Besser G, Renner B, Mueller CA. Olfactory-related Quality of Life Adjustments in Smell Loss during the Coronavirus-19 Pandemic. Am J Rhinol Allergy. 2022;36(2):253–260. doi:10.1177/19458924211053118

15. Mattos JL, Schlosser RJ, Storck KA, Soler ZM. Understanding the relationship between olfactory-specific quality of life, objective olfactory loss, and patient factors in chronic rhinosinusitis. Int Forum Allergy Rhinol. 2017;7(7):734–740. doi:10.1002/alr.21940

16. Malaty J, Malaty IAC. Smell and taste disorders in primary care. Am Fam Physician. 2013;88(12):852–859.

17. Risso D, Drayna D, Morini G. Alteration, Reduction and Taste Loss: Main Causes and Potential Implications on Dietary Habits. Nutrients. 2020;12(11):3284. doi:10.3390/nu12113284

18. Sanderson AL, Burns JP. Clinical Documentation for Intensivists: The Impact of Diagnosis Documentation. Crit Care Med. 2020;48(4):579–587. doi:10.1097/CCM.0000000000004200

19. Hosseini N, Mostafavi SM, Zendehdel K, Eslami S. Factors affecting clinicians’ adherence to principles of diagnosis documentation: A concept mapping approach for improved decision-making. Health Inf Manag J Health Inf Manag Assoc Aust. 2022;51(3):149–158. doi:10.1177/1833358321991362

20. Momin SR, Lorenz RR, Lamarre ED. Effect of a Documentation Improvement Program for an Academic Otolaryngology Practice. JAMA Otolaryngol Neck Surg. 2016;142(6):533–537. doi:10.1001/jamaoto.2016.0194

21. Olmstead J. Understanding the importance of diagnosis coding. Nurse Pract. 2018;43(10):8. doi:10.1097/01.NPR.0000544998.00017.de

22. Pinto JM, Wroblewski KE, Kern DW, Schumm LP, McClintock MK. Olfactory dysfunction predicts 5-year mortality in older adults. PloS One. 2014;9(10):e107541. doi:10.1371/journal.pone.0107541

23. Holdoway A. Nutritional management of patients during and after COVID-19 illness. Br J Community Nurs. 2020;25(Sup8):S6–S10. doi:10.12968/bjcn.2020.25.Sup8.S6

24. Hummel T, Liu DT, Müller CA, Stuck BA, Welge-Lüssen A, Hähner A. Olfactory Dysfunction: Etiology, Diagnosis, and Treatment. Dtsch Arzteblatt Int. 2023;120(9):146-154. doi:10.3238/arztebl.m2022.0411

25. Beecher K, St John JA, Chehrehasa F. Factors that modulate olfactory dysfunction. Neural Regen Res. 2018;13(7):1151–1155. doi:10.4103/1673-5374.235018

26. Bang Y, Lim J, Choi HJ. Recent advances in the pathology of prodromal non-motor symptoms olfactory deficit and depression in Parkinson’s disease: clues to early diagnosis and effective treatment. Arch Pharm Res. 2021;44(6):588–604. doi:10.1007/s12272-021-01337-3

27. Doty RL. Chapter 15 - Psychophysical testing of smell and taste function. In: Doty RL, ed. Handbook of Clinical Neurology. Vol 164. Smell and Taste. Elsevier; 2019:229-246. doi:10.1016/B978-0-444-63855-7.00015-0

28. Rombaux P, Collet S, Martinage S, et al. Olfactory testing in clinical practice. B-ENT. 2009;5 Suppl 13:39–51.

29. Frequently Asked Questions | Smell and Taste Center | Perelman School of Medicine at the University of Pennsylvania. Accessed May 26, 2023. https://www.med.upenn.edu/smellandtastecenter/faq.html

30. Saraswathula A, Schlosser RJ, Rowan NR. Reimbursement for Olfactory Testing: A Growing Need at a Unique Time in History. JAMA Otolaryngol Neck Surg. Published online May 11, 2023. doi:10.1001/jamaoto.2023.0837

31. CPT® overview and code approval. American Medical Association. Accessed May 18, 2023. https://www.ama-assn.org/practice-management/cpt/cpt-overview-and-code-approval

32. Shih MC, Soler ZM, Germroth M, Snyder J, Nguyen SA, Schlosser RJ. Comparison of validated psychophysical olfactory tests and olfactory-specific quality of life. Int Forum Allergy Rhinol. 2022;12(11):1428–1431. doi:10.1002/alr.23012

33. Brief Smell Identification Test® (B-SIT®). Sensonics International. Accessed May 18, 2023. https://sensonics.com/product/brief-smell-identification-test-b-sit-version-cross-cultural-smell-identification-test/

34. Smell Identification Test^TM^ (UPSIT®). Sensonics International. Accessed May 18, 2023. https://sensonics.com/product/smell-identification-test/

35. Dalton P, Doty RL, Murphy C, et al. Olfactory assessment using the NIH Toolbox. Neurology. 2013;80(11 Suppl 3):S32-S36. doi:10.1212/WNL.0b013e3182872eb4

36. Odor Identification Test. NIH Toolbox. Accessed June 27, 2023. https://www.nihtoolbox.org/test/odor-identification-test/

37. Hummel T, Sekinger B, Wolf SR, Pauli E, Kobal G. “Sniffin” sticks’: olfactory performance assessed by the combined testing of odor identification, odor discrimination and olfactory threshold. Chem Senses. 1997;22(1):39–52. doi:10.1093/chemse/22.1.39

38. Rumeau C, Nguyen DT, Jankowski R. How to assess olfactory performance with the Sniffin’ Sticks test®. Eur Ann Otorhinolaryngol Head Neck Dis. 2016;133(3):203–206. doi:10.1016/j.anorl.2015.08.004

39. Sniffin’ Sticks - US Neurologicals. Accessed May 18, 2023. https://usneurologicals.com/Item/ST_SniffinSticks

40. Cingoz ID, Kizmazoglu C, Guvenc G, Sayin M, Imre A, Yuceer N. Evaluation of the Olfactory Function With the “Sniffin’ Sticks” Test After Endoscopic Transsphenoidal Pituitary Surgery. J Craniofac Surg. 2018;29(4):1002–1005. doi:10.1097/SCS.0000000000004398

41. Parma V, Hannum ME, O’Leary M, et al. SCENTinel 1.0: development of a rapid test to screen for smell loss. Chem Senses. Published online March 27, 2021. doi:10.1093/chemse/bjab012

42. Hunter SR, Hannum ME, Pellegrino R, et al. Proof-of-concept: SCENTinel 1.1 rapidly discriminates COVID-19-related olfactory disorders. Chem Senses. 2023;48:bjad002. doi:10.1093/chemse/bjad002

43. Weir EM, Hannum ME, Reed DR, et al. The Adaptive Olfactory Measure of Threshold (ArOMa-T): a rapid test of olfactory function. Chem Senses. 2022;47:bjac036. doi:10.1093/chemse/bjac036

44. Chang MT, Patel ZM. Novel Therapies in Olfactory Disorders. Curr Otorhinolaryngol Rep. 2022;10(4):427–432. doi:10.1007/s40136-022-00436-z

45. Ahmed K, Wang TT, Ashrafian H, Layer GT, Darzi A, Athanasiou T. The effectiveness of continuing medical education for specialist recertification. Can Urol Assoc J. 2013;7(7-8):266–272. doi:10.5489/cuaj.378

46. IMPORTANCE of CME. American Association of Continuing Medical Education®. Accessed May 18, 2023. https://aacmet.org/cme/importance-of-cme/

47. Chandra A, Dervenoulas G, Politis M, Alzheimer’s Disease Neuroimaging Initiative. Magnetic resonance imaging in Alzheimer’s disease and mild cognitive impairment. J Neurol. 2019;266(6):1293–1302. doi:10.1007/s00415-018-9016-3

48. Chung MS, Choi WR, Jeong HY, Lee JH, Kim JH. MR Imaging–Based Evaluations of Olfactory Bulb Atrophy in Patients with Olfactory Dysfunction. Am J Neuroradiol. 2018;39(3):532–537. doi:10.3174/ajnr.A5491

49. Pagano G, Niccolini F, Politis M. Imaging in Parkinson’s disease. Clin Med Lond Engl. 2016;16(4):371–375. doi:10.7861/clinmedicine.16-4-371

