## Supplementary material for "Patient Insights into the Diagnosis of Smell and Taste Disorders in the United States": PCORI Survey Questions

Jefferson Health
HOME OF SIDNEY KIMMEL MEDICAL COLLEGE

MONELL CENTER
ADVANCING DISCOVERY IN TASTE AND SMELL

### Screening Questions

You are being asked to participate in a survey that is led by the Monell Chemical Senses Center, Smell and Taste Association of North America (STANA), and Jefferson Health and funded by a grant from the Patient-Centered Outcomes Research Institute (PCORI).

The survey is limited to residents of the United States of America. The survey should take about 15 minutes to complete. Upon completion, you can opt to enter and provide your contact information for a raffle for one of twenty-five $50 gift cards.

Your survey responses will be anonymous and no personally identifying information will be associated with the survey. There are no known risks or discomforts associated with answering these questions. Your participation is completely voluntary and if you choose not to participate, to discontinue participation in this study, or skip some questions there will be no penalty or loss of benefits, and this will not affect your future interactions with Monell, STANA or Jefferson Health.

The purpose of this research survey is to understand the impact of smell or taste dysfunction from COVID-19 and other causes on a variety of everyday experiences and to gain a better understanding of any barriers in diagnosis or treatment for smell and taste loss. Monell,

STANA and Jefferson Health have partnered to develop a prioritized research agenda that creates a framework for future patient-centered research, including updating a 2018 White Paper outlining research needed to advance treatments.

Your answers will be kept confidential. The data collected from this survey will be used for research purposes only, will be analyzed and stored by the study team at Monell, STANA and Jefferson, and may be used in the future to answer other research questions. If you have any questions, concerns, or complaints regarding your participation in this survey, please contact: the Principal Investigator, Dr. Pamela Dalton, at 267-519-4810 or, or the Co-Investigator, Katie Boateng, at. If you would like to speak with someone other than the study sponsors, you may contact the Office of Regulatory Affairs with any question, concerns, or complaints at the University of Pennsylvania by calling 215-573-2540.

Please state whether you consent to participate in this study:

- ○ Yes, I consent to participate in this study
- ○ No, I do not consent to participate in this study

Relationship to smell and/or taste disorder

- ○ Patient
- ○ Family or caregiver of a patient (not a parent)
- ○ Parent of a patient
- ○ None of the above

Country of Residence

- United States of America
- Other

Throughout this survey, you are going to be asked to rate your senses of taste and smell.

Taste is defined as the ability to perceive chemicals on the tongue as sweet, salty, sour, bitter, or umami/savory (for example cooked mushrooms or soy sauce).

Smell is defined as the ability to perceive odors or scents.

**Demographic Information**

Gender

- Male
- Female
- Non-binary / third gender
- Prefer not to say
- ______ Other (fill in the blank below)

Age

- Under 18
- 18 - 24
- 25 - 39
- 40-60
- 60 or older

Do you consider yourself:

- ○ White
- ○ Black or African American
- ○ American Indian or Alaska Native
- ○ Asian
- ○ Native Hawaiian or Pacific Islander
- ○ Prefer not to answer
- ○ ________ Other (fill in the blank below)

Are you of Spanish or Hispanic origin, such as Latin American, Mexican, Puerto Rican, Cuban, or Dominican?

- ○ Yes, of Hispanic origin
- ○ No, not of Hispanic origin
- ○ Prefer not to answer

In which state do you currently reside?

[dropdown]

What is your zip code?

________________

What type of health insurance do you have?

- I do not have any health insurance
- Individual health insurance policy (purchased directly from a health insurer or through a health insurance exchange)
- Employer or other group health insurance
- Military/TRICARE
- Medicare
- Medicaid
- ______ Other (fill in the blank below)

Do you see a healthcare provider for preventative visits or regular check-ups?

- Yes
- No

What type of healthcare provider do you see for preventative visits or regular check-ups?

- Primary Care Clinician (MD, DO, NP, PA, etc.)
- Otolaryngologist (Ear Nose Throat - ENT) Specialist
- Nurse practitioner
- Neurologist
- Chiropractor
- Dentist
- Acupuncturist
- Taste and Smell Clinic
- Allergist
- Oncologist

- ☐ OB/Gynecologist
- ☐ Eye doctor (ophthalmologist) or optometrist
- ☐ ________ Other (fill in the blank below)

### Disease Experience/Diagnosis

Which of the following have you ever experienced? Check all that apply.

- ☐ Complete smell loss (anosmia)
- ☐ Reduced smell loss or partial loss of smell (hyposmia)
- ☐ Distorted smells (parosmia)
- ☐ Phantom smells (phantosmia)
- ☐ Complete taste loss (ageusia)
- ☐ Distorted taste (dysgeusia/parageusia)
- ☐ Reduced taste loss or partial loss of taste (hypogeusia)
- ☐ None

Is your smell/taste dysfunction due to Covid-19?

- ○ Yes
- ○ No
- ○ I don't know

**What would you like your healthcare provider to know about your condition?**

Is your smell/taste dysfunction officially documented in your medical record?

- ☐ Yes
- ☐ No
- ☐ I don't know

Which of the following is officially documented in your medical record?

- ○ Complete smell loss (anosmia)
- ○ Reduced smell loss or partial loss of smell (hyposmia)
- ○ Distorted smells (parosmia)
- ○ Phantom smells (phantosmia)
- ○ Complete taste loss (ageusia)
- ○ Distorted taste (dysgeusia/parageusia)
- ○ Reduced taste loss or partial loss of taste (hypogeusia)
- ○ I don't know

What do you think caused your parosmia?

- ○ Cold/flu
- ○ Allergies

- ○ Trauma to the nose or face
- ○ Stroke or other neurologic event
- ○ Migraine
- ○ Virus (not Covid-19)
- ○ Covid-19 (virus)
- ○ ______ Other (fill in the blank below)

What types of things trigger your parosmia?

______

**What do you think caused your anosmia?**

- ○ Traumatic brain injury (TBI)
- ○ Congenital
- ○ Viral (not related to Covid-19)
- ○ Viral (Covid-19)
- ○ Aging
- ○ Idiopathic (cause is unknown)
- ○ ______ Other (fill in the blank below)

Diagnosis status

- ○ I do not have an official diagnosis but I am certain I have one of these conditions
- ○ I do not have an official diagnosis and I am unsure whether I have one of these conditions

To the best of your knowledge, when did you first **experience** your symptoms of smell/taste dysfunction?
Select a Date (feel free to approximate):

| | Month | Year |
|---|---|---|
| Please Select: | | |

To the best of your knowledge, when were you first **diagnosed** with your symptoms of smell/taste dysfunction? If not applicable (for example, you do not have an official diagnosis) please leave blank.
Select a Date (feel free to approximate):

| | Month | Year |
|---|---|---|
| Please Select: | | |

To the best of your knowledge, when did your smell/taste dysfunction **end**? If not applicable (for example, you're still experiencing it) please leave blank.
Select a Date (feel free to approximate):

| | Month | Year |
|---|---|---|
| Please Select: | | |

How would you have rated your sense of smell prior to the onset of your symptoms? (Smell is defined as the ability to perceive odors or scents.)

- Extremely bad

- Somewhat bad
- Neither good nor bad
- Somewhat good
- Extremely good

How would you have rated your sense of taste prior to the onset of your symptoms? (Taste is defined as the ability to perceive chemicals on the tongue as salty, sweet, sour, bitter, or umami/savory [example: cooked mushrooms or soy sauce].)

- Extremely bad
- Somewhat bad
- Neither good nor bad
- Somewhat good
- Extremely good

Once you realized you were experiencing smell/taste dysfunction, which of the following did you do?

- [ ] Visit a healthcare provider
- [ ] Look for information online
- [ ] Talk to friends and family
- [ ] ________ Other (fill in the blank below)

If you sought out information on social media, which of the following did you seek out? Check all that apply.

- ☐ Facebook
- ☐ Twitter
- ☐ LinkedIn
- ☐ TikTok
- ☐ Instagram
- ☐ ____________ Other (fill in the blank below)

If you looked online for information, which of the following places did you use? Select all that apply.

- ☐ Academic websites (Mayo, Hopkins, etc.)
- ☐ General search (Google, Bing, Yahoo, etc.)
- ☐ ____________ Other (fill in the blank below)

**Medical Provider**

Did you see a healthcare provider for your symptoms?

- ○ No
- ○ Yes

Why not?

What type of healthcare provider did you visit? Check all that apply.

- ☐ Family practitioner
- ☐ Otolaryngologist (Ear Nose Throat - ENT) Specialist
- ☐ Nurse practitioner
- ☐ Neurologist
- ☐ Chiropractor
- ☐ Dentist
- ☐ Acupuncturist
- ☐ Taste and Smell Clinic
- ☐ Allergist
- ☐ Oncologist
- ☐ ____________ Other (fill in the blank below)

### Follow up Questions to Q42

Did the ${lm://Field/1} do any of the following? Check all that apply.

- ☐ Provide written materials
- ☐ Provide a referral to another specialist
- ☐ Provide social support

- ☐ None of the above
- ☐ ______ Other (fill in the blank below)

How knowledgeable was the ${lm://Field/1} about smell/taste dysfunction?

- ◯ Not knowledgeable at all
- ◯ Slightly knowledgeable
- ◯ Moderately knowledgeable
- ◯ Very knowledgeable
- ◯ Extremely knowledgeable

How supported (listened to and believed) by the ${lm://Field/1} did you feel <u>during</u> your visit?

- ◯ Very supported
- ◯ Supported
- ◯ Somewhat supported
- ◯ Not at all supported

How supported (listened to and believed) by the ${lm://Field/1} did you feel <u>after</u> your visit?

- ◯ Very supported
- ◯ Supported
- ◯ Somewhat supported
- ◯ Not at all supported

What was your confidence level that the ${lm://Field/1} could help you manage or treat your symptoms?

- ○ Very confident
- ○ Confident
- ○ Somewhat confident
- ○ Not at all confident

### Diagnosis

If you had a difficult time getting a diagnosis, are there any specific reasons you think played a role in that difficulty? Check all that apply.

- ☐ I do not feel like I was listened to and/or believed
- ☐ Lack of health insurance
- ☐ Lack of provider knowledge about diagnostic testing
- ☐ The cost of testing was prohibitive
- ☐ Difficulty getting access to a specialist to get a diagnosis
- ☐ My symptoms were attributed to another disease that I already had a diagnosis for
- ☐ I did not have a hard time getting diagnosed
- ☐ ________ Other (fill in the blank below)

What tests or diagnostic procedures did you undergo? Check all that apply.

- ☐ Smell test (UPSIT, Sniffin' Sticks, etc.)
- ☐ Taste test

- ☐ MRI
- ☐ CT
- ☐ Nasal endoscopy
- ☐ Questionnaire (Example: SNOT-22)
- ☐ Medical history
- ☐ ____________ Other (fill in the blank below)
- ☐ None of the above

How many healthcare providers did you see for the problem before you got a diagnosis?

- ○ 1
- ○ 2
- ○ 3
- ○ more than 3
- ○ I do not have a diagnosis
- ○ ____________ Other (fill in the blank below)

What treatment options did your healthcare provider recommend? Check all that apply.

- ☐ Nasal steroids
- ☐ Oral steroids
- ☐ Zinc
- ☐ Nasal rinse
- ☐ Smell training
- ☐ Alpha lipoic acid
- ☐ Theophylline

- ☐ Omega-3
- ☐ PRP (platelet rich plasma)
- ☐ Participating in a clinical trial
- ☐ ______ Other (fill in the blank below)
- ☐ None of the above

**Follow up to Q46 : What Treament**

Do you/did you face any of the following barriers or difficulties in accessing your selected treatment: (${lm://Field/1})?

- ☐ Cost
- ☐ Insurance coverage
- ☐ Physical proximity to appointments for treatment
- ☐ Amount of time the treatment takes
- ☐ Side effects from the treatment
- ☐ ______ Other (fill in the blank below)
- ☐ I do not face any barriers or difficulties

If (${lm://Field/1}) was used, please rate how effective it has been.

- ○ Not effective at all
- ○ Slightly effective
- ○ Moderately effective
- ○ Very effective
- ○ Extremely effective

### Condition's Impact on Quality of Life

To what degree (if any) has your smell/taste dysfunction affected the following lifestyle functions for you?

| | Not at all | Somewhat | Mildly or slightly | Moderately | Very | Extremely |
|---|---|---|---|---|---|---|
| Nutrition (diet, weight loss, weight gain) | ○ | ○ | ○ | ○ | ○ | ○ |
| Eating habits (quantity, preferences, other changes) | ○ | ○ | ○ | ○ | ○ | ○ |
| Cooking ( level of enjoyment, frequency, ability) | ○ | ○ | ○ | ○ | ○ | ○ |
| Food choices (color, texture, temperatures) | ○ | ○ | ○ | ○ | ○ | ○ |
| Appetite | ○ | ○ | ○ | ○ | ○ | ○ |
| Relationship and bonding with others (significant others, family members, children, etc. | ○ | ○ | ○ | ○ | ○ | ○ |
| How much you enjoy dining out with others | ○ | ○ | ○ | ○ | ○ | ○ |
| Work | ○ | ○ | ○ | ○ | ○ | ○ |
| Mental health (depression, anxiety, grief) | ○ | ○ | ○ | ○ | ○ | ○ |
| Memory | ○ | ○ | ○ | ○ | ○ | ○ |
| Pleasure/Joy | ○ | ○ | ○ | ○ | ○ | ○ |
| Safety | ○ | ○ | ○ | ○ | ○ | ○ |
| Sex and intimacy | ○ | ○ | ○ | ○ | ○ | ○ |

| | Not at all | Somewhat | Mildly or slightly | Moderately | Very | Extremely |
|---|---|---|---|---|---|---|
| Personal hygiene habits | ○ | ○ | ○ | ○ | ○ | ○ |
| How much your worry/focus on hygiene | ○ | ○ | ○ | ○ | ○ | ○ |
| Other (fill in the blank below) | ○ | ○ | ○ | ○ | ○ | ○ |

Is there anything you wish others understood about the impact of smell/taste dysfunction on your quality of life?

- ○ Yes
- ○ No

What do you wish others understood about the impact of smell/taste dysfunction on your quality of life?

Have you specifically sought treatment for your mental health in dealing with your smell/taste dysfunction?

- ○ Yes
- ○ No

Do you think your smell/taste dysfunction is a disability?

- Yes
- No

Please expand on why you think your smell/taste dysfunction is a disability.

Please expand on why you think your smell/taste dysfunction is **not** a disability.

Of all the possibilities for support for people with smell/taste dysfunction, please rate the following items on level of importance to you.

| | Not at all important | Slightly important | Moderately important | Very important | Most Important |
|---|---|---|---|---|---|
| More public awareness about the condition | O | O | O | O | O |

| | Not at all important | Slightly important | Moderately important | Very important | Most Important |
|---|---|---|---|---|---|
| More medical community knowledge about the condition | ○ | ○ | ○ | ○ | ○ |
| Effective treatments for the condition | ○ | ○ | ○ | ○ | ○ |
| Better diagnostic tools for the condition | ○ | ○ | ○ | ○ | ○ |
| Supportive devices (electronic noses, gas detection, etc.) | ○ | ○ | ○ | ○ | ○ |
| Lifestyle strategies for compensating | ○ | ○ | ○ | ○ | ○ |
| Mental health support | ○ | ○ | ○ | ○ | ○ |
| Other (fill in the blank below) ____ | ○ | ○ | ○ | ○ | ○ |
| Other (fill in the blank below) ____ | ○ | ○ | ○ | ○ | ○ |
| Other (fill in the blank below) ____ | ○ | ○ | ○ | ○ | ○ |

### Information/Systems for Support

What are your primary sources of information about smell/taste dysfunction? Check all that apply.

- ☐ The provider I see for my smell and/or taste loss or distortion
- ☐ Friends/Family
- ☐ Other people with my disorder

- ☐ Social media sites (Twitter, Facebook, TikTok, Instagram, etc.)
- ☐ Search engines like Google
- ☐ Academic/scientific organization websites
- ☐ Patient advocacy organizations
- ☐ Magazines, books, or newsletters
- ☐ Television or radio
- ☐ In-person events or educational classes
- ☐ I do not seek any information
- ☐ ____________ Other (fill in the blank below)

What are your primary sources of <u>support</u> for dealing with or coping with your smell/taste dysfunction? Check all that apply.

- ☐ The provider I see for my smell and/or taste loss or distortion
- ☐ Friends/Family
- ☐ Other people with my disorder
- ☐ Social media sites (Twitter, Facebook, TikTok, Instagram, etc.)
- ☐ Search engines like Google
- ☐ Academic/scientific organization websites
- ☐ Patient advocacy organizations
- ☐ Magazines, books, or newsletters
- ☐ Television or radio
- ☐ In-person events or educational classes
- ☐ I do not seek any support
- ☐ ____________ Other (fill in the blank below)

### Looking Ahead

If a new treatment were to be developed for smell/taste dysfunction, what are the top three factors that would impact whether or not you would try it?

- ☐ Effectiveness of the drug or treatment for treating the most troublesome symptoms
- ☐ Cost or insurance barriers
- ☐ Method of administration (injection, infusion, oral, topical)
- ☐ Required frequency of treatments
- ☐ Required frequency administering doses
- ☐ Psychological effect (fear, nervousness) of the therapy
- ☐ Common but not serious side effects (headache, nausea, etc.)
- ☐ Uncommon but serious side effects (risk of cancer, anaphylaxis, etc.)
- ☐ Travel distance to clinic/provider to receive the therapy
- ☐ Duration of treatment (days, weeks, months to complete)
- ☐ __________ Other (fill in the blank below)

Would you be willing to participate in research for smell/taste dysfunction?

- ○ Yes
- ○ No

Of the following types of research, rate how likely you would be to participate

| | Extremely unlikely | Somewhat unlikely | Neither likely nor unlikely | Somewhat likely | Extremely likely |
|---|---|---|---|---|---|

| | Extremely unlikely | Somewhat unlikely | Neither likely nor unlikely | Somewhat likely | Extremely likely |
|---|---|---|---|---|---|
| Clinical trial for a new pharmaceutical treatment | ○ | ○ | ○ | ○ | ○ |
| Research project for a surgical treatment | ○ | ○ | ○ | ○ | ○ |
| Research project for a non-pharmaceutical or non-surgical treatment (example: a study about the effectiveness of smell training) | ○ | ○ | ○ | ○ | ○ |
| Research focused on causes or risk factors for smell/taste disorders or mechanisms for smell and taste regeneration (example: genetic testing to identify genes involved in smell loss or regeneration) | ○ | ○ | ○ | ○ | ○ |

Why not?

In your opinion, what is the most effective way to inform patients about advances in research and treatments for smell/taste dysfunction?

When you think about the future, do any of the following items concern you? Check all that apply.

- ☐ Your sense of smell and/or taste will only recover partially
- ☐ Your sense of smell and/or taste will not recover at all
- ☐ Your sense of smell and/or taste may be lost or distorted again
- ☐ None of the above
- ☐ __________ Other (fill in the blank below)

Do you feel like you have been treated differently in any of the following environments? Check all that apply.

- ☐ Workplace
- ☐ Family
- ☐ Social setting
- ☐ __________ Other (fill in the blank below)
- ☐ I do not feel like I've been treated differently

How do people react when they hear about your smell/taste dysfunction?

What do you think people find most difficult to understand about your smell/taste dysfunction?

Are there any other aspects of your smell/taste dysfunction you would like to share with us?

Powered by Qualtrics
